# Memory and executive functioning show differential associations with scam susceptibility in middle to older adulthood

**DOI:** 10.64898/2026.10.04.26364617

**Authors:** Cassidy P. Molinare, Belinda Y. Zhang, Mark Sanderson-Cimino, Melanie Leguizamon, Emma Oyen, Daisy T. Noriega-Makarskyy, Jenna Axelrod, Aaron C. Lim, Camdyn Wu, Maanya Agarwal, Laura Mosqueda, S. Duke Han

## Abstract

**Objectives:** The rising prevalence of financial exploitation among older adults has prompted research on the cognitive mechanisms underlying financial decision making in later life. Prior work has demonstrated a link between cognitive impairment and financial vulnerability. The present study examined the unique contributions of composite measures of memory and executive functioning to scam susceptibility in middle-aged to older adults without dementia.

**Methods:** One hundred and fifty-one participants (age= 68.2 ± 8.0 years, 72.2% female, years of education= 16.4 ± 2.2) completed standard neuropsychological measures from the Uniform Data Set (UDS), Version 3, and the California Verbal Learning Test, Second Edition, and a self-report scam susceptibility measure. Psychometrically robust composite scores, the UDS-M+ and UDS3-EF, were computed to represent memory and executive functioning domains. Linear regression models examined independent associations between UDS-M+ and UDS3-EF performance and scam susceptibility. Nested model comparisons evaluated unique contributions of the UDS-M+ and UDS3-EF when included in a joint model.

**Results:** Lower UDS-M+ (b= -0.25, 95% CI [-0.45, -0.04], *p*= .018) and lower UDS3-EF (b= - 0.33, 95% CI [-0.54, -0.12], *p*= .002) scores were independently associated with greater scam susceptibility after adjusting for age, sex, years of education, and income. However, only UDS3-EF remained significant in a joint model. Post hoc analyses revealed that only memory was associated with scam susceptibility in middle-aged adults whereas only executive functioning was significant in older adults.

**Conclusions:** These findings highlight the importance of monitoring memory and executive functioning across middle to older adulthood when considering susceptibility to scams.

**Statement of Research Significance:** *Research Question(s) or Topic(s):* This study evaluated the contributions of memory and executive functioning to scam susceptibility in middle-aged to older adults without dementia using cognitive composite scores and whether these associations differed across middle-aged and older adult groups.

*Main Findings:* After adjusting for age, sex, education, and income, memory (b= -0.25, 95% CI [-0.45, -0.04], *p*= .018) and executive functioning (b= -0.33, 95% CI [-0.54, -0.12], *p*= .002) each demonstrated an independent inverse association with scam susceptibility. In a joint model, only executive functioning was associated with scam susceptibility. When comparing age groups, only memory was significant in middle age while only executive functioning was significant in older adults.

*Study Contributions:* These results extend prior research by clarifying the unique contributions of memory and executive functioning to financial vulnerability and suggesting there may be differences in the cognitive processes used for financial decision making across stages of older adulthood.

## 1. Introduction

Financial exploitation in older age has been previously defined by the Center for Disease Control & Prevention as the illegal, unauthorized, or improper use of an older adult’s money, benefits, property, or assets for the benefit of another person, and is considered one of the most common types of elder abuse (CDC, 2024). Financial abuse can occur in various forms including theft, scams, or fraud, and is often characterized by deception, manipulation, or breach of trust (Wood & Lichtenberg, 2017). According to the FBI’s latest Annual Internet Crime Report, older adults lost $7.7 billion to cybercrime and scams in 2025, a 61.4% increase from the $4.8 billion reported in 2024 (FBI Internet Crime Complaint Center, 2025, 2026). However, the American Association of Retired Persons (AARP) estimates that after accounting for unreported cases, older adults lose approximately $28.3 billion annually due to financial exploitation (Gunther, 2023). The magnitude of these financial losses reflects the severity and scope of financial exploitation among older adults and highlights the need to better understand the factors that contribute to suboptimal financial decision making in later life.

Older adults are thought to be most vulnerable to financial exploitation due to wealth accumulation (Maynard et al., 2025), lower social connectedness (Noriega-Makarskyy et al., 2025), and age-related changes in cognitive functioning (Agarwal et al., 2009; Lachs & Han, 2015; Tymula et al., 2013). However, recent evidence has suggested that vulnerability is not uniform across all older adults. Individuals who exhibit cognitive deficits consistent with the preclinical stages of Alzheimer’s disease (AD) may be particularly susceptible to poor financial decision making. For example, Weissberger et al. (2023) found that a neuropsychological profile consistent with early AD, characterized in part by lower performances on verbal memory measures, was associated with greater blind giving, or a greater willingness to give away money to a stranger rather than keeping it for oneself. Additionally, several studies have demonstrated that older adults with mild cognitive impairment (MCI) exhibit greater scam susceptibility than cognitively unimpaired individuals (Han et al., 2015b, 2015a). Evidence from longitudinal work has further shown that poorer financial decision making and lower scam awareness are associated with more rapid rates of cognitive decline and a greater risk of incident AD (Boyle et al., 2012, 2013, 2019; Stewart et al., 2019). Taken together, these findings indicate that suboptimal financial decision making and greater scam susceptibility may coincide with changes in cognitive functioning that occur in early AD.

Alzheimer’s disease is characterized by a prolonged preclinical period during which neuropathology, such as amyloid-β burden and tau deposition, develops years before the onset of overt clinical symptoms (Braak & Braak, 1991; Jack et al., 2013; Sperling et al., 2011). One of the earliest cognitive changes observed during this period is a decline in memory functioning. In particular, lower performance on episodic memory and delayed recall measures is considered a hallmark feature of the disease and can be detected several years prior to clinical diagnosis (Bäckman et al., 2005; Small et al., 2000). Consistent with evidence linking suboptimal financial decision making to an increased risk of progression to MCI and AD, prior work examining the cognitive correlates of financial decision-making measures has identified consistent associations between episodic memory and financial vulnerability. Specifically, across studies of older adults with and without cognitive impairment, lower episodic memory performance has been associated with greater susceptibility to scams (Han et al., 2015a; James et al., 2014), greater perceived financial exploitation vulnerability (Lim et al., 2024), greater blind giving (Weissberger et al., 2022), and lower financial literacy, a supportive construct for financial decision making (Boyle et al., 2013; Gamble et al., 2015; Weissberger et al., 2023). As such, greater financial vulnerability may reflect a disruption in memory functioning that occurs at the earliest stages of AD.

While there is compelling evidence linking memory deficits to suboptimal financial decision making, decision making has traditionally been associated with executive functioning as it relies on processes like planning, reasoning, cognitive flexibility, and impulse control. A substantial body of literature has supported this association, often using the Iowa Gambling Task (Bechara et al., 1994), a paradigm designed to mimic real-life decisions, as the primary decision-making measure (see Toplak et al., 2010 for a review). These studies have shown that poorer decision making is associated with worse performance on standard measures of executive functioning (e.g., Trail Making, Wisconsin Card Sorting), reflecting demands on cognitive flexibility and inhibition of impulsive choices (Brand et al., 2006, 2007; Denburg et al., 2005). Consistently, studies that used alternate measures of decision making, such as the Adult Decision-Making Competence battery (De Bruin et al., 2007), also found associations between decision making performance and executive abilities, including working memory and verbal fluency (Del Missier et al., 2010, 2012; Rosi et al., 2019).

Research to date has demonstrated links between financial decision-making measures and performance on both individual neuropsychological tests and cognitive z-score composites (James et al., 2014), implicating specific aspects of memory and executive functioning in later life financial vulnerability. However, prior work examining broader executive functioning composite scores with memory composite scores is limited. Furthermore, it remains unclear whether deficits in memory and executive functioning independently contribute to older adults’ susceptibility to scams and fraud, and whether each domain accounts for unique variance when considered jointly. The present study sought to address this gap by leveraging robust memory and executive functioning composite scores to examine the associations between memory, executive functioning, and scam susceptibility in a sample of older adults without dementia. Employing item response theory (IRT)-derived composite scores, which incorporate various traditional neuropsychological measures into domain-specific factor scores, enables a more comprehensive understanding of these mechanisms as they relate to scam susceptibility. We hypothesized that lower memory and executive functioning performance would each be associated with greater scam susceptibility in separate and joint models.

## 2. Methods

### 2.1 Participants

Participants included community-dwelling older adults from the Finance, Cognition, and Default Network in Aging (FCDNA) study at the University of Southern California. They were recruited from community outreach events, local senior centers, and the Alzheimer’s Prevention Registry (Langbaum et al., 2020). Eligible participants were aged 50 years or older, fluent in English, and free from dementia (scored ≥ 21 on a modified 5-minute Montreal Cognitive Assessment administered during their phone screening; Wong et al., 2015). Exclusion criteria included a history of significant neurological or psychiatric illness, alcohol use disorder in the last five years, or any history of other substance abuse. During their study visit, participants completed a questionnaire in which they self-reported their date of birth, sex (binarily coded as male or female), years of education, race and ethnicity, and income. Income was classified as participants’ total combined family income during the past 12 months, including “money from jobs, net income from business, farm or rent, pensions, dividends, interest, Social Security payments, and any other monetary income received by members of the family who are at 15 years of age or older”. Multiple-choice response options included: 1 – Less than $5,000, 2 – $5,000 to $7,499, 3 – $7,500 to $9,999, 4 – $10,000 to $12,499, 5 – $12,500 to $14,999, $15,000 to $19,999, 7 – $20,000 to $24,999, 8 – $25,000 to $29,999, 9 – $30,000 to $34,999, 10 – $35,000 to $39,999, 11 – $40,000 to $49,999, 12 – $50,000 to $59,999, 13 – $60,000 to $74,999, 14 – $75,000 to $99,999, 15 – $100,000 to $149,999, 16 – $150,000 or more. This income question was drawn from the standard demographic variables collected by the Understanding America Study (UAS), a large, nationally representative internet panel study on the daily lives of U.S. families and individuals that is maintained by the Center for Economic and Social Research (CESR) at the University of Southern California (available at https://uasdata.usc.edu/index.php; Alattar et al., 2018; Kapteyn et al., 2024). All study procedures were completed in accordance with the Helsinki Declaration and were approved by the University of Southern California institutional Review Board, and all participants provided their written consent upon enrollment.

### 2.2 Measure

#### 2.2.1 Neuropsychological assessments

The Mini-Mental State Examination was administered to participants as a measure of global cognition (Folstein et al., 1975). All participants completed the California Verbal Learning Test, Second Edition (CVLT-II; Delis et al., 2000) and the neuropsychological test battery from the Uniform Data Set, Version 3 (UDS-3; Weintraub et al., 2018), including the Craft Story 21, Benson Complex Figure, Trail Making Test Parts A and B, Multilingual Naming Test (MINT), phonemic fluency, and semantic fluency measures.

#### 2.2.2 Scam susceptibility

Participants completed a 5-item self-report measure of scam susceptibility (Boyle et al., 2012; Han et al., 2015a, 2021; James et al., 2014), with items derived from findings from the AARP and the Financial Industry Regulatory Authority Risk Meter, a measure of risky financial decision making that is often used in finance studies (AARP, 1999; Financial Industry Regulatory Authority, 2013). Items on this measure included “I answer the phone whenever it rings, even if I do not know who is calling” and “If something sounds too good to be true, it usually is”. Participants rated their agreement with each item using a 7-point Likert scale (1 = strongly agree, 7 = strongly disagree). Total scores were calculated as an average of responses, with some items reverse scored. Total scores ranged between 1 and 7, with higher scores reflecting greater scam susceptibility.

### 2.3 Composite Score Creation

The executive functioning (UDS3-EF) and memory (UDS-M+) composite scores were computed in the present study using the published scoring (Staffaroni et al., 2020) and harmonization (Sanderson-Cimino et al., 2026) pipelines. The UDS3-EF and UDS-M+ use IRT-based estimation methods to calculate latent factor scores (Gross et al., 2023; Mukherjee et al., 2022), which may be more statistically robust than traditional composite score creation methods (e.g., averaged z-scores) as they account for item difficulty and discrimination (Chapman & Chapman, 1978; Crane et al., 2023), and were designed for use in datasets where the subset of completed items differed by participants (Sanderson-Cimino et al., 2026; Staffaroni et al., 2020).

In the present study, the UDS3-EF was calculated based on the following measures of executive functioning from the UDS-3: Digit Span Backwards (total correct), Trail Making Test Parts A and B (correct lines per minute), phonemic fluency (F and L words total correct), and semantic fluency (animals and vegetables total correct). In the present study, all component subtests from the UDS-3 were available, and only three participants did not complete Trail Making Test Part B. For these three participants, the UDS3-EF was estimated without the Trail Making Test Part B subtest. The UDS-M+ was derived from the following tasks: the CVLT-II (immediate recall, long delay free and cued recall, recognition hits, and recognition false positives); Craft Story 21 immediate and delayed recall, Benson Complex Figure delayed recall and recognition. (Chapman & Chapman, 1978; Crane et al., 2023).

### 2.4 Statistical Analyses

All analyses were conducted using R, version 4.5.2. Separate linear regression models were used to examine the associations of executive functioning (UDS3-EF scores) and memory (UDS-M+ scores) with scam susceptibility, adjusting for age, sex, years of education, and income. For each model, we evaluated the proportion of explained variance (*R*^2^) to determine the extent to which executive functioning and memory independently accounted for variability in scam susceptibility. We also estimated an additional linear regression model including both UDS3-EF and UDS-M+ scores as predictors. The unique contributions of executive functioning and memory were quantified using changes in explained variance (Δ*R*^2^), calculated as the difference between the *R*^2^ of the full model and the *R*^2^ of the corresponding reduced model. This approach estimates the variance uniquely attributable to each predictor after accounting for the other. Each domain’s unique contribution was evaluated using analysis of variance (ANOVA) to compare nested models, assessing whether including memory significantly improved model fit beyond the executive functioning-only model and whether including executive functioning significantly improved model fit beyond the memory-only model. All models controlled for participants’ age, sex, and years of education, and income.

Post hoc analyses applied a median age split (<69 and ≥69 years old) to determine whether significant associations between memory, executive functioning, and scam susceptibility were consistent across middle-aged and older adult participants. A median split was used as a data-driven approach to classifying middle-aged and older adult subgroups within our sample, given its wide age range.

## 3. Results

### 3.1 Sample Characteristics

The full sample included 151 older adults without dementia. Participants’ age ranged from 51 to 87 years old (*M* age = 68.2, *SD* = 8.0), with 74 individuals in the middle-aged group (<69 years) and 77 individuals in the older adult group (≥69 years). The sample was 72.2% female and, on average, highly educated (*M* years of education = 16.4, *SD* = 2.2). Participants were 65.6% White, 17.2% Asian, 8.6% Black or African American, 5.3% Mixed Race, 2.6% Other, and 0.7% Unknown. See Table 1 for complete demographic information and descriptive statistics. Group differences in sample characteristics between the middle-aged and older adult age groups were evaluated using chi-square tests for categorical variables, independent samples *t*-tests for continuous variables, and Wilcoxon rank-sum tests for ordinal variables (Table 1). There was a significant group difference in the racial breakdown of each group (*p* < 0.001), which was driven by differences in the proportion of individuals who identified as a mixed race or “Other”.

**Table 1.** Sample characteristics and descriptive statistics across the full sample and between middle-aged and older adult groups.

| **Characteristic** | **Overall,**<br>**N=151** | **<69 years old,**<br>**(n=74)** | **≥69 years old,**<br>**(n=77)** | **<69 vs. ≥69,**<br>**p-value** |
| --- | --- | --- | --- | --- |
| Age, Mean (SD) | 68.2 (8.0) | 61.6 (4.71) | 74.6 (4.4) | <.001 |
| Sex, n (%) Female | 109 (72.2) | 53 (71.6%) | 56 (72.7%) | 1 |
| Years of Education, Mean (SD) | 16.4 (2.2) | 16.5 (2.1) | 16.2 (2.4) | .421 |
| Race, n (%) |  |  |  | <.001 |
| White | 99 (65.6) | 48 (64.9%) | 51 (66.2%) |  |
| Asian | 26 (17.2) | 14 (18.9%) | 12 (15.6%) |  |
| Black or African American | 13 (8.6) | 6 (8.1%) | 7 (9.1%) |  |
| Mixed Race | 8 (5.3) | 6 (8.1%) | 2 (2.6%) |  |
| Other | 4 (2.6) | 0 (0%) | 4 (5.2%) |  |
| Unknown | 1 (0.7) | 0 (0%) | 1 (1.3%) |  |
| MMSE, Mean (SD) | 28.2 (1.7) | 28.4 (1.7) | 28.0 (1.6) | .208 |
| Income, Median | 14 | 14 | 14 | .666 |
| UDS3-EF, Mean (SD) | 0.08 (0.77) | 0.36 (0.76) | -0.19 (0.69) | <.001 |
| UDS-M+, Mean (SD) | 0.25 (0.46) | 0.32 (0.47) | 0.19 (0.43) | .234 |
| Scam Susceptibility, Mean (SD) | 1.96 (0.88) | 1.96 (0.79) | 1.96 (0.96) | .991 |

*Note.* Income was measured using a 16-point scale (13: \$60,000 to \$74,999; 14: \$75,000 to \$99,999). Group differences between the middle-aged and older adult groups were examined using chi-square tests for categorical variables, independent samples *t*-tests for continuous variables, and Wilcoxon rank-sum tests for ordinal variables. MMSE = Mini-Mental State Examination; UDS3-EF = executive functioning composite; UDS-M+ = memory composite.

### 3.2 Primary Analyses

Lower performance on the UDS-M+ was associated with greater scam susceptibility (b = -0.25, 95% CI [-0.45, -0.04], *p* = .018). Lower performance on the UDS3-EF was also associated with greater scam susceptibility (b = -0.33, 95% CI [-0.54, -0.12], *p* = .002). All models controlled for age, sex, years of education, and income. After adjusting for covariates, the model including UDS-M+ performance explained 8.2% of the variance in participants’ scam susceptibility scores (*R*^2^ = 0.082), whereas the model including UDS3-EF performance and the same covariates explained 10.6% of the variance in scam susceptibility (*R*^2^ = 0.106).

In a full model that included both the UDS-M+ and UDS3-EF as predictors, UDS3-EF performance was inversely associated with scam susceptibility scores (b = -0.27, 95% CI [-0.50, -0.05], *p* = .015), while UDS-M+ performance was not (b = -0.16, 95% CI [-0.38, 0.05], *p* = .136). The full model explained 11.9% of the variance in scam susceptibility scores (*R*^2^ = 0.119). The UDS3-EF accounted for 2.3% more unique variance in scam susceptibility than the UDS-M+ and significantly improved model fit beyond the UDS-M+ (*F*(1,144) = 6.02, *p* = .015, Δ*R^2^*= 0.037), whereas the UDS-M+ did not improve model fit beyond the UDS3-EF (*F*(1,144) = 2.25, *p* = .136, Δ*R^2^*= 0.014).

### 3.3 Post hoc analyses

Separate linear regression analyses were conducted within middle-aged and older adult age groups to examine the associations between UDS-M+ performance, UDS3-EF performance, and scam susceptibility, adjusting for age, sex, years of education, and income. Lower UDS-M+ performance was associated with greater scam susceptibility in the middle-aged group (b = -0.42, 95% CI [-0.66, -0.18], *p* = .001), but not in the older age group (b = -0.08, 95% CI [-0.42, 0.26], *p* = .639). By contrast, lower UDS3-EF performance was associated with greater scam susceptibility in the older age group (b = -0.44, 95% CI [-0.79, -0.10], *p* = .02), but not in the middle-aged group (b = -0.21, 95% CI [-0.47, 0.05], *p* = .105).

In the middle-aged group, the full model that included both the UDS-M+ and UDS3-EF as predictors showed an inverse association between UDS-M+ performance and scam susceptibility (b = -0.39, 95% CI [-0.64, -0.14], *p* = 0.003), while UDS3-EF performance was not significant (b = -0.08, 95% CI [-0.34, 0.18], *p* = 0.520). This full model explained 29.1% of the variance in participants’ scam susceptibility scores (*R*^2^ = 0.291). The UDS-M+ accounted for 9.7% more unique variance in scam susceptibility than the UDS3-EF and significantly improved model fit beyond the UDS3-EF (F(1,67) = 9.57, *p* = .003, Δ*R^2^*= 0.101), whereas the UDS3-EF did not improve model fit beyond the UDS-M+ (F(1,67) = 0.42, *p* = .520, Δ*R^2^*= 0.004).

Conversely, in the older age group, the full model including both the UDS-M+ and UDS3-EF showed an inverse association between UDS3-EF performance and scam susceptibility (b = -0.46, 95% CI [-0.83, -0.10], *p* = 0.014), while UDS-M+ performance was not significant (b = 0.06, 95% CI [-0.29, 0.41], *p* = 0.723). Here, the full model explained 16.5% of the variance in scam susceptibility scores (*R*^2^ = 0.165). The UDS3-EF accounted for 7.6% more unique variance in scam susceptibility than the UDS-M+ and significantly improved model fit beyond the UDS-M+ (*F*(1,70) = 6.39, *p* = .014, Δ*R^2^*= 0.076), whereas the UDS-M+ did not improve model fit beyond the UDS3-EF (*F*(1,70) = 0.13, *p* = .723, Δ*R^2^*= 0.002).

**Figure 1.**
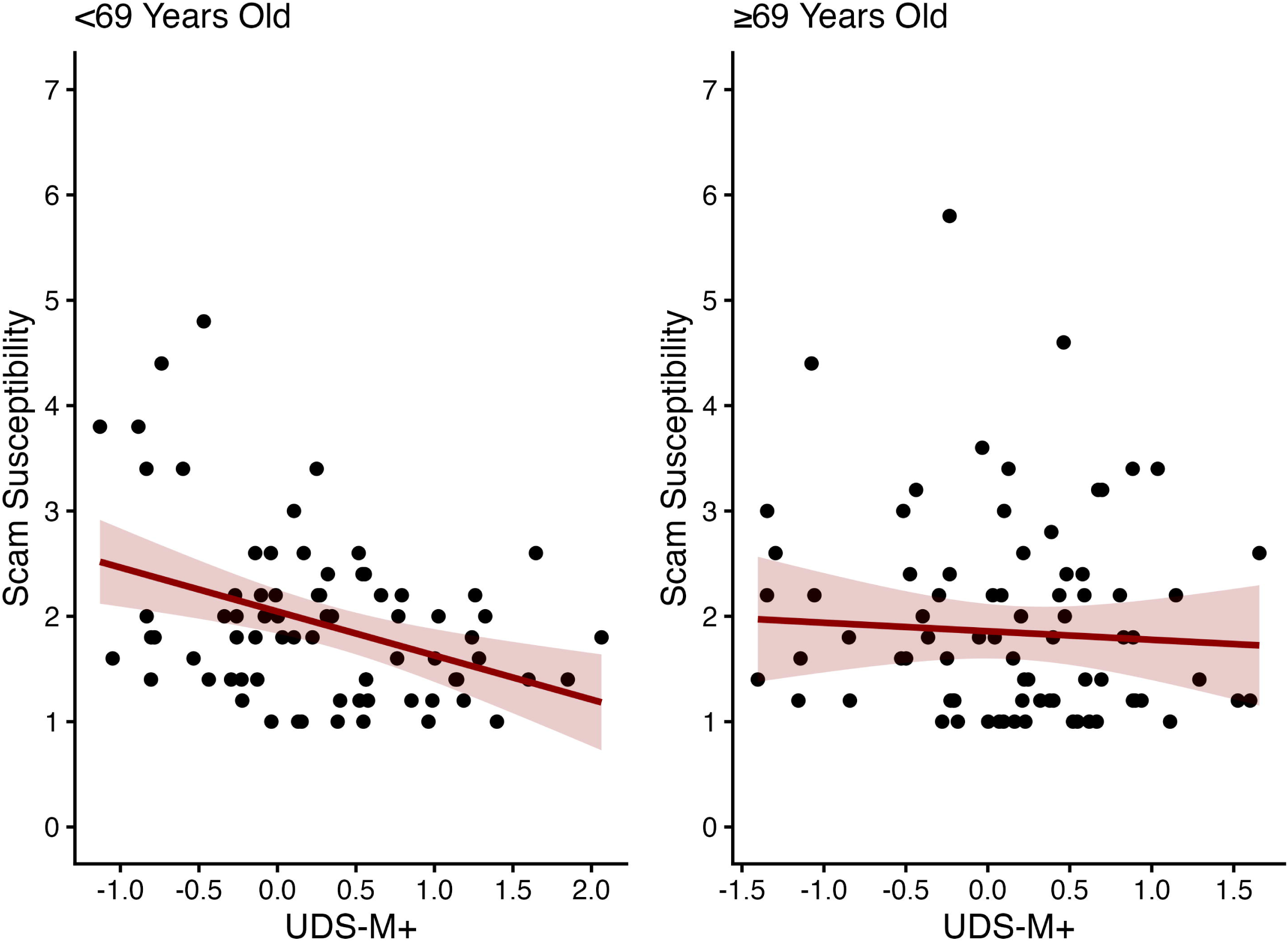
Associations between memory performance (UDS-M+) and scam susceptibility by age group *Note.* Points represent observed data. Lines represent predicted values from linear regression models adjusted for age, sex, years of education, and income. Shaded areas indicate 95% confidence intervals around the predicted values.

**Figure 2.**
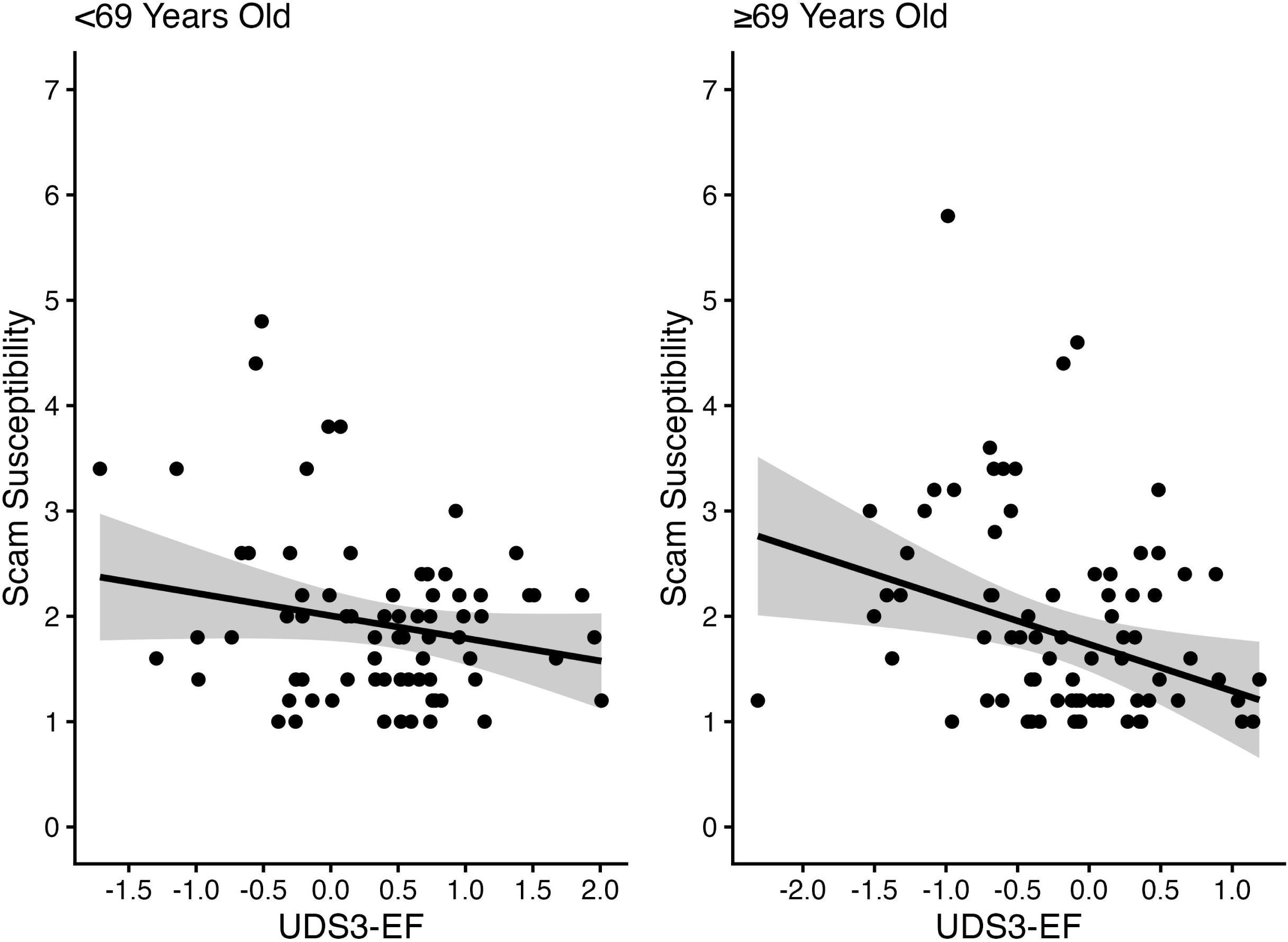
Associations between executive functioning performance (UDS3-EF) and scam susceptibility by age group *Note.* Points represent observed data. Lines represent predicted values from linear regression models adjusted for age, sex, years of education, and income. Shaded areas indicate 95% confidence intervals around the predicted values.

## 4. Discussion

The purpose of the present study was to investigate the associations between memory, executive functioning, and scam susceptibility in middle-aged and older adults without dementia. Consistent with our hypothesis, we found that lower memory performance was associated with greater scam susceptibility after adjusting for age, sex, years of education, and income. Similarly, lower executive functioning performance was associated with greater scam susceptibility after adjusting for the same covariates. However, after examining the unique contributions of the memory and executive functioning composites to variance in scam susceptibility in a joint model, we found that only executive functioning remained a significant predictor. Nested model comparisons further demonstrated that adding the executive functioning composite to a model with the memory composite improved model fit, while adding the memory composite to a model with the executive functioning composite did not improve model fit. In post hoc analyses that compared middle-aged (<69 years) and older adult age groups (≥69 years), we found that memory, but not executive functioning, was inversely associated with scam susceptibility in the middle-aged group, while executive functioning, but not memory, was associated with scam susceptibility in the older adult group. Our findings extend prior work linking cognitive functioning to scam susceptibility in older adults by clarifying the distinct roles of memory and executive functioning and how their unique contributions vary across middle-aged and older adult age groups.

The independent associations between memory, executive functioning, and scam susceptibility align with existing literature in which scam susceptibility has been consistently associated with MCI and progression to AD (Boyle et al., 2012, 2019; Han et al., 2015a; James et al., 2014). Notably, both memory and executive functioning are known to decline during the preclinical stages of AD (Albert, 1996; Clark et al., 2012; Guarino et al., 2019; Jahn, 2013; Storandt, 2008), suggesting that increased susceptibility to scams may reflect cognitive changes associated with early neurodegenerative processes. Building on earlier findings linking specific memory abilities and executive functions to financial decision making and vulnerability (Boyle et al., 2013; Gamble et al., 2015; James et al., 2014; Lim et al., 2024; Molinare et al., under review; Weissberger et al., 2022, 2023), our results showed that broader, IRT-derived composite measures of memory and executive functioning are also associated with scam susceptibility.

However, beyond their independent associations, we found that when including memory and executive functioning performance in the same model, only executive functioning remained a significant contributor to variance in scam susceptibility. This finding suggests that while memory may play a role in scam susceptibility, only executive functioning is a significant unique predictor. The importance of executive functioning to financial vulnerability is consistent with previous research linking executive dysfunction to impairments in instrumental activities of daily living, including financial management (Marshall et al., 2011). Further, previously observed associations between cognitive impairment or dementia and poorer financial decision making may be more attributable to declines in executive functioning that occur alongside memory decline, as prior studies did not directly compare the unique contributions of these domains.

Executive functioning is thought to encompass three core functions: mental set-shifting or cognitive flexibility, inhibitory control, and working memory (Lehto, 1996; Lehto et al., 2003; Miyake et al., 2000), which together are essential for financial decision making. Cognitive flexibility and working memory are critical as they allow individuals to evaluate different courses of action and adapt when they discover information that might indicate fraud. While not directly assessed in the UDS3-EF, inhibitory control may be particularly relevant for regulating an impulse to donate money or share sensitive financial information that may arise when a stranger “in need” asks for money or if an opportunity sounds “too good to be true”. Beyond these core functions, other executive abilities such as planning, reasoning, and problem-solving are vital for making logical decisions that align with one’s goals (Collins & Koechlin, 2012; Cristofori et al., 2019; Lezak et al., 2004). These functions allow individuals to think critically, anticipate potential challenges, and consider the consequences of their decisions, which are necessary processes for combating scam attempts and making more optimal financial decisions.

Results from our post hoc analyses provide preliminary evidence that middle-aged and older adults may rely on distinct cognitive processes for decision making, whereby middle-aged adults may engage in more memory-related processes while older adults might rely more on executive functioning abilities. Although we are unable to determine the mechanisms underlying these differences in the present study, one theoretical framework that may align with these findings is the Selective Optimization with Compensation (SOC) theory, which suggests that older adults adapt to age-related changes in cognitive functioning by selectively prioritizing specific goals, optimizing performance in preserved domains, and compensating for declines in other domains through alternative strategies (B. B. Baltes & Dickson, 2001; P. B. Baltes & Baltes, 1990). Within this framework, as memory declines in later life due to age or neurodegenerative processes, older adults may shift toward greater use of executive functions to support their decision making. While executive functioning may play a more consistent role in financial decision making across older adulthood, individuals with more intact memory abilities, particularly middle-aged individuals, may continue to rely on processes like prospection. An additional theoretical framework that may help explain these findings is the Socioemotional Selectivity Theory (SST), which posits that as individuals perceive their future time horizons as more limited, they increasingly prioritize present-oriented goals, particularly emotional meaning and satisfaction, whereas individuals with longer perceived time horizons typically exhibit more future-oriented thinking (Carstensen, 2021; Carstensen et al., 1999; Lang & Carstensen, 2002). These age-related differences in time perspectives and motivations may shape the cognitive processes that individuals employ during financial decision making. Middle-aged adults may be more likely to engage in prospective memory processes during financial decision making, considering how the consequences of their actions correspond with their long-term goals, whereas older adults may focus on salient information in the present context and goals with emotional significance. In the context of financial vulnerability, older adults’ financial decisions may be more strongly influenced by the urgent and emotional appeals that scam perpetrators seek to elicit, with less relative consideration of future outcomes. Such decisions may therefore depend on processes that support the evaluation and regulation of immediate responses, such as working memory, inhibitory control, and reasoning, rather than episodic memory.

There are several limitations to this work that should be considered. First, our study sample was predominately White, female, and highly educated. Given the lack of demographic representation in our sample, the generalizability of our findings is limited. As such, there is a need to replicate these analyses in a larger, population-based sample. Second, the present study was cross-sectional, so we cannot infer causal relationships. Future longitudinal work may examine whether memory and executive functioning performances at baseline are associated with subsequent increases in vulnerability to scams or fraud. Third, the cognitive composite scores are limited by the measures used to create them, as the UDS-3 neuropsychological battery only includes traditional measures intended for use in Alzheimer’s disease research centers.

Future research may aim to replicate these findings by creating composite scores that incorporate newer cognitive measures assessing aspects of executive functioning and memory not detected by traditional measures. For example, digital measures such as the Face Name Associative Memory Exam (Jutten et al., 2021; Rentz et al., 2011, 2023), a paired associative memory task, and digital batteries like the Delis-Kaplan Executive Function System (D-KEFS) Advanced (Delis & Kaplan, 2025), which assesses higher-order executive functions, may help capture additional nuances in memory and executive functioning. Similarly, our scam susceptibility measure relies on self-report responses, which may be subject to inaccuracies due to social desirability or limited insight into one’s own behavior, particularly in those with declining memory or executive functioning. Incorporating more objective measures of scam and fraud victimization in future studies would improve ecological validity.

Despite these limitations, the results of the present study offer greater insight into the associations among memory, executive functioning, and scam susceptibility across middle-aged and older adults. By clarifying the cognitive mechanisms underlying scam susceptibility in later life, these findings may help identify early indicators of financial vulnerability and support caregivers or family members in recognizing when intervention may be indicated. Furthermore, these results emphasize the importance of assessing and monitoring cognitive changes in older adults, particularly executive functioning, as it may be useful for identifying individuals at risk and informing efforts to protect older adults from financial abuse.

## Acknowledgements

We thank the participants for their time and contribution to this work.

## Funding Statement

This work was supported by the National Institute on Aging (R01AG068166 and K24AG081325 to SDH). Some participants from the present study were recruited with the help of the Alzheimer’s Prevention Registry. The Alzheimer’s Prevention Registry is supported by a grant from the National Institute on Aging (R01AG063954). The Alzheimer’s Prevention Registry has been supported by the Alzheimer’s Association, Banner Alzheimer’s Foundation, Flinn Foundation, Geoffrey Beene Gives Back Alzheimer’s Initiative, GHR Foundation, and the state of Arizona (Arizona Alzheimer’s Consortium).

## Competing Interests

The authors declare that there are no conflicts of interest.

## Data Availability Statement

The data used in the present study has been deidentified and made publicly available on Zenodo: https://doi.org/10.5281/zenodo.23092391

## Notes

### Competing Interest Statement

The authors have declared no competing interest.

### Author Declarations

The University of Southern California Institutional Review Board gave ethical approval for this work.

